# A streamlined approach to rapidly detect SARS-CoV-2 infection, avoiding RNA purification

**DOI:** 10.1101/2020.04.06.20054114

**Authors:** Stefania Marzinotto, Catia Mio, Adriana Cifù, Roberto Verardo, Corrado Pipan, Claudio Schneider, Francesco Curcio

**Affiliations:** Department of Laboratory Medicine, ASU FC, Udine, Italy; Department of Medicine (DAME), University of Udine, Udine, Italy; CIB-Consorzio Interuniversitario Biotecnologie (Interuniversity Consortium for Biotechnologies), Padriciano 99 34012 Trieste

## Abstract

In the current pandemic, the presence of SARS-CoV-2 RNA in samples by nucleic acid (NA) molecular analysis is the only method available to diagnose COVID-19 disease and to assess patients’ infectiveness. Recently, the demand for laboratory reagents has greatly increased; in particular, there is a worldwide shortage of RNA extraction kits. Here, we describe a fast, simple and inexpensive method for the detection of SARS CoV-2 RNA, which includes a pretreatment with Proteinase K and a heating-cooling cycle before the amplification. This method bypasses the RNA extraction step; it leads to a higher amount of available viral RNA compared to the automated extraction methods, and generates the same profile in the subsequent amplification phase.

## Introduction

Since the outbreak of the newly appeared severe acute respiratory syndrome coronavirus-2 (SARS-CoV-2) caused coronavirus disease (COVID-19), the demonstration of the virus presence by nasopharyngeal swabs and subsequent molecular analysis of viral Nucleic Acid (NA) has been the preferred diagnostic method. Several NA automated analytical molecular systems are available, which, although having the advantage of obtaining a pure product of the highest quality, are expensive and greatly lengthen the analysis procedures. More importantly, recently the ability of manufacturers of reagents for molecular diagnostics to meet the increasing demand has progressively diminished. In particular, the main limitation is represented by the shortage of NA extraction kits. Since the demonstration of the virus in the samples is the only way to really indicate the patient’ infectiveness, this shortage risks to greatly impair our ability to limit the spreading of the pandemic with tragic consequences. In addition, even when reagents are available, NA isolation represents a significant part of the analytical protocol; eliminating this step will increase the productivity of laboratories and decrease the cost of testing.

To solve the above-mentioned issues, we developed a procedure for the treatment of nasopharyngeal swab (n=500) collected from patients with suspected SARS-CoV-2 infection which eliminates the need for the RNA extraction step.

## Materials and Methods

### Automated RNA extraction from nasopharyngeal swab

For nasopharyngeal swab collection, UTM^®^ tubes (COPAN Diagnostics) were used, i.e. plastic, screw cap tubes that maintains organism viability for 48 hours at room temperature or less. Those are FDA-approved systems suitable for collection, transportation and long-term storage (−20°C) of clinical specimens. 200 µL of medium from 3mL of UTM^®^ tube were used for automated RNA extraction with the ELITe InGenius^®^ SP200 System (ELITechGroup), following manufacturer’s instructions. Samples were eluted in 100µL elution buffer. Ethical approval was obtained from the Medical Research Ethics Committee of the Region Friuli Venezia Giulia, Italy (Consent CEUR-2020-Os-033).

### RNA isolation from nasopharyngeal swab

In a 96-well plate, 100µL of medium from 3mL of UTM^®^ tube were added to 10µL of 30mg/mL Proteinase K from Tritirachium album (Sigma-Merck, catalog number P2308) in Hanks’ Balanced Salt Solution (HBSS) w/ calcium and magnesium and w/o phenol red (Sigma-Merck, catalog number 55037C). The plate was heated for 15min at 55°C, denatured for 5min at 98°C and then placed for 2min at 4°C.

### Quantitative Reverse Transcription Polymerase Chain Reaction (RT-qPCR)

LightMix^®^ Modular SARS and Wuhan CoV E-gene (Roche, catalog number 53-0776-96) was used to compare the amplification of the *E-gene* in nasopharyngeal swab samples in which the RNA was purified by an automated system with those treated by the method developed by us. Briefly, 4µL Roche Master, 0.5µL Reagent mix (containing specific primers and probe following Corman et al. 2020)^1^, 0.1µL RT enzyme, 5 µL sample/positive control and nuclease-free water were mixed to a total volume of 15µL. RT-qPCR was performed by the LightCycler^®^ 480 II Instrument (Roche) and absolute quantification was assessed by the LightCycler^®^ 480 II System (Roche).

### One-Step Reverse Transcription-Droplet digital Polymerase Chain Reaction (RT-ddPCR)

5’ 6-FAM/3’ BHQ-1^®^-conjugated *E-gene*^1^ (Sigma Merck) was used for viral load assessment by the One-Step RT-ddPCR Advanced Kit for Probes (Bio-Rad, catalog number 186-4021). Briefly, 5µL of ddPCR™ Supermix for Probes (No dUTP), 900 nM primers and 250 nM probes, 15mM DTT, 20U/µL Reverse Transcriptase, 5 µL sample and nuclease-free water were mixed and brought to a total volume of 20 µL. Samples were mixed with Droplet Generator Oil for Probes (Bio-Rad, catalog number 1863005) and droplets were generated on the automated droplet generator QX200™ Droplet Generator (Bio-Rad). PCR amplification was performed by the Veriti^®^ Thermal Cycler (ThermoFisher Scientific) with annealing at 55°C and standard thermal cycling conditions. Droplets were read on the QX200™ Droplet Reader (Bio-Rad) and reactions with less than 10’000 droplets were repeated. Data were analyzed using the QuantaSoft™ 1.7.4 Software (Bio-Rad).

### Statical analysis

The statistical analyses were performed with GraphPad Prism 6.0. Shapiro-Wilk test was performed to assess the normality of data distribution. Data are reported as mean ± SD. Pearson’s linear regression was performed to assess correlation between measurements. Paired Student’s t test was performed to compare the two populations. *p<0.05, **p<0.01, ***p<0.001, **** p<0.0001

## Results and Discussion

A great number of nucleic acid extraction kits are used to prepare viral samples for analysis. In early 2020, Italy faced the exponential outbreak of SARS-CoV-2 infection originated in China and since declared a pandemic by the WHO, which highly impacted on the healthcare systems around the World and was responsible for tens of thousands of deaths. Molecular testing for SARS-CoV-2^2^ is currently the best available approach for a correct diagnosis and to establish the infective potential of patients. RNA extraction from nasopharyngeal swabs of patients affected by COVID-19 has become a bottleneck in diagnostic procedures due to the enormous quantity of samples to be daily processed and to the shortage in extraction kit availability. In many instances, the shortage has induced the limitation of testing only to patients with symptoms (often non-specific) but in this way many asymptomatic or paucisymptomatic individuals^3,4^ have not been tested and have become a major vehicle by which the infection has spread, first in China^5^ and then in the rest of the World. To overcome this issue, we set up an in-house protocol to pretreat swab samples before performing RT-qPCR.

We compared results of the amplifications from automatedly extracted viral RNA with the swab-derived material treated as described in methods.

As shown in Figure 1, Panel A, both samples displayed the same amplification profile with minimal differences. To further validate our data, we employed a technique with higher sensitivity, the droplet digital PCR (ddPCR). Using samples from the automated extraction and from our method, we found the same viral copies (i.e. 13000 copies/5µL) in both, showing that our in-house pretreatment protocol is a reliable and rapid method to assess SARS-CoV-2 infection directly from nasopharyngeal swabs (Fig. 1; Panel B). Furthermore, since the automated diagnostic extraction system used in this study requires 200µL swab-derived material as template, while our method only needs 100µL of material, we were able to detect twice as much of the viral genome for each µL of sample.

**Figure 1.**
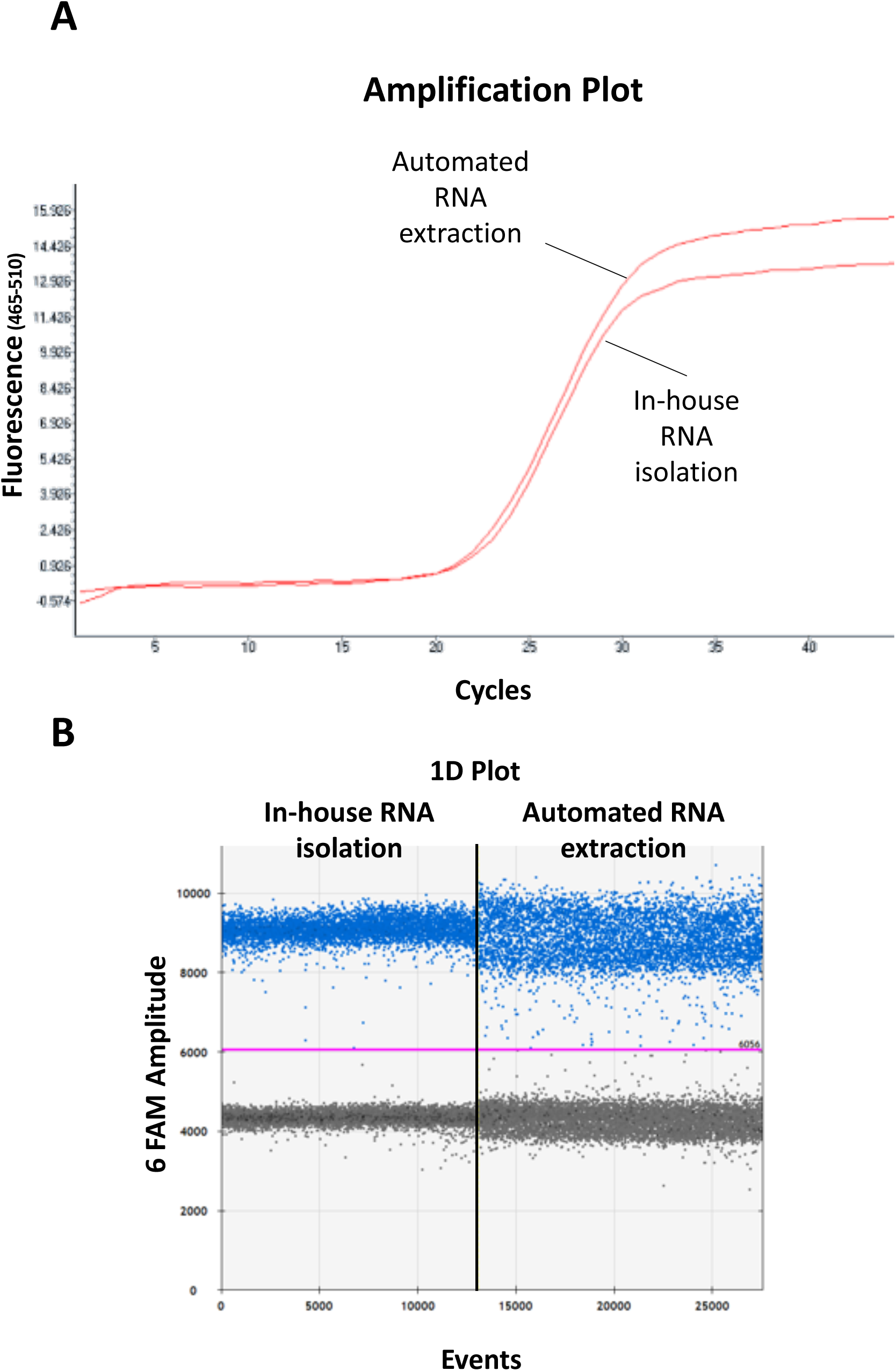
Comparison between automated RNA extraction and in-house RNA isolation. Patient’s sample processed either with automated extraction or the in-house optimized procedure. A, the same amplification efficiency (average 22.5 Cp) was observed despite the diverse amount of swab-derived medium used for extraction/isolation (200µL automated extraction vs 100µL custom protocol) in RT-qPCR. B, the same viral load (average 13000 copies) was observed in both samples by RT-ddPCR. In the figure, one representative amplification profile is shown.

Moreover, to assess correlation between our in-house RNA isolation method and the automated RNA extraction, linear regression was performed using RT-qPCR amplification data from 17 samples. Shapiro-Wilk’s normality test was performed and, as shown in Figure 2, Pearson r between the two groups was 0.9276 (CI 95% 0,8066-0,9740, p < 0.0001) suggesting a high correlation between methodologies.

**Figure 2.**
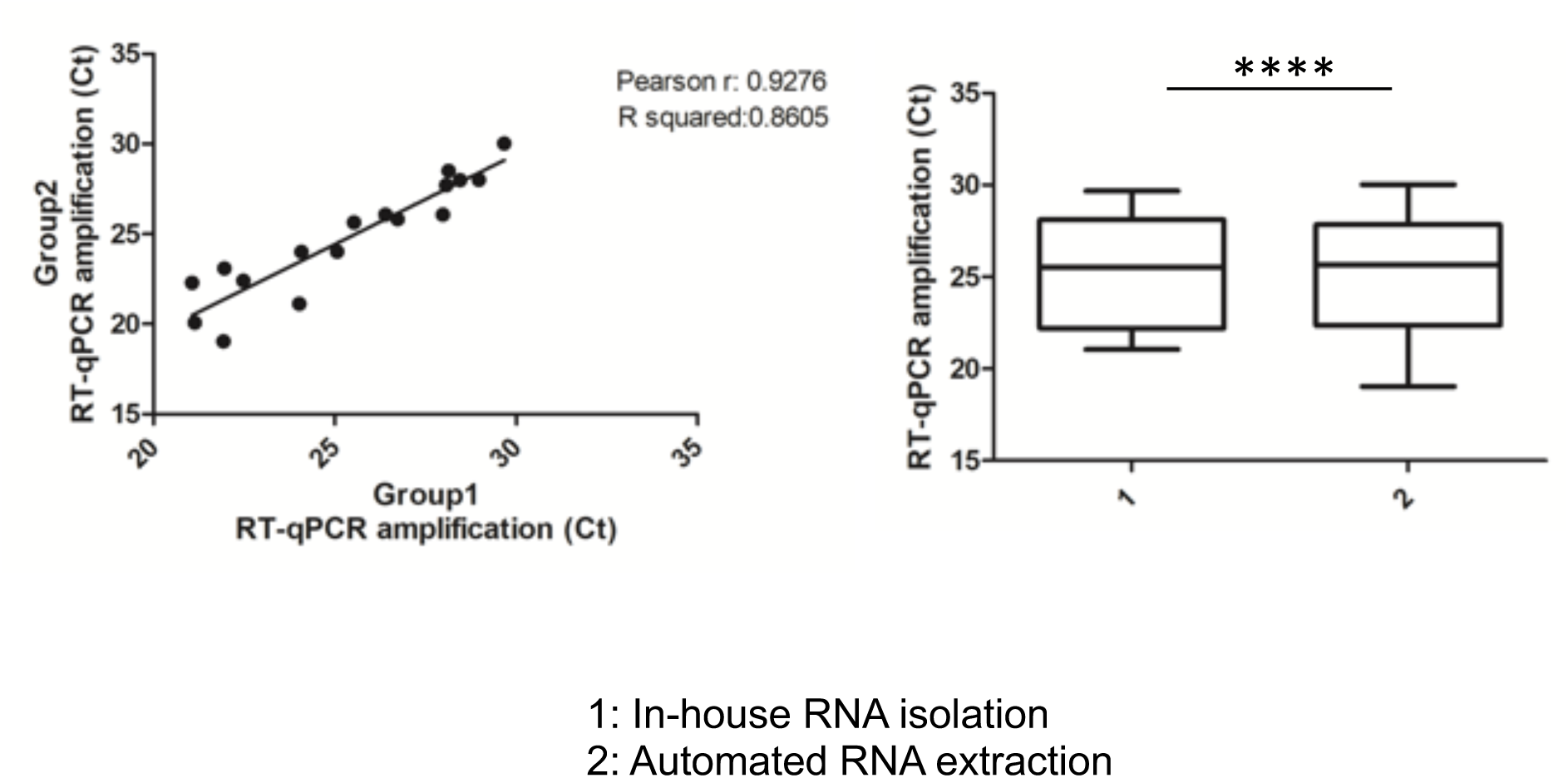
Correlation between in-house RNA isolation and automated RNA extraction. Scatter plot with linear regression (left panel) between the two methodologies tested. Black line represents the best-fit regression line. Box plot (right panel) representing the mean ± SD of N=17 samples treated with the two methodologies and amplified by RT-qPCR. ****p<0.0001.

Considering the performance of our pretreatment protocol in generating a higher amount of viral RNA compared to the automated one, we tested its sensitivity in assessing viral copies with the two amplification methods (RT-qPCR and RT-ddPCR). We performed a 5-fold serial dilution of the same sample used in Figure 1 and assessed amplification curves in RT-qPCR. As shown in Figure 3, panel A, we were able to evaluate viral copies even in dilutions as high as 1:3125. To effectively assess how many actual viral copies our protocol is able to detect, we used the same approach using RT-ddPCR. As shown in Figure 3, panel B, our in-house protocol is able to detect as few as 10 SARS-CoV-2 copies directly from 5µL nasopharyngeal swab-derived material.

**Figure 3.**
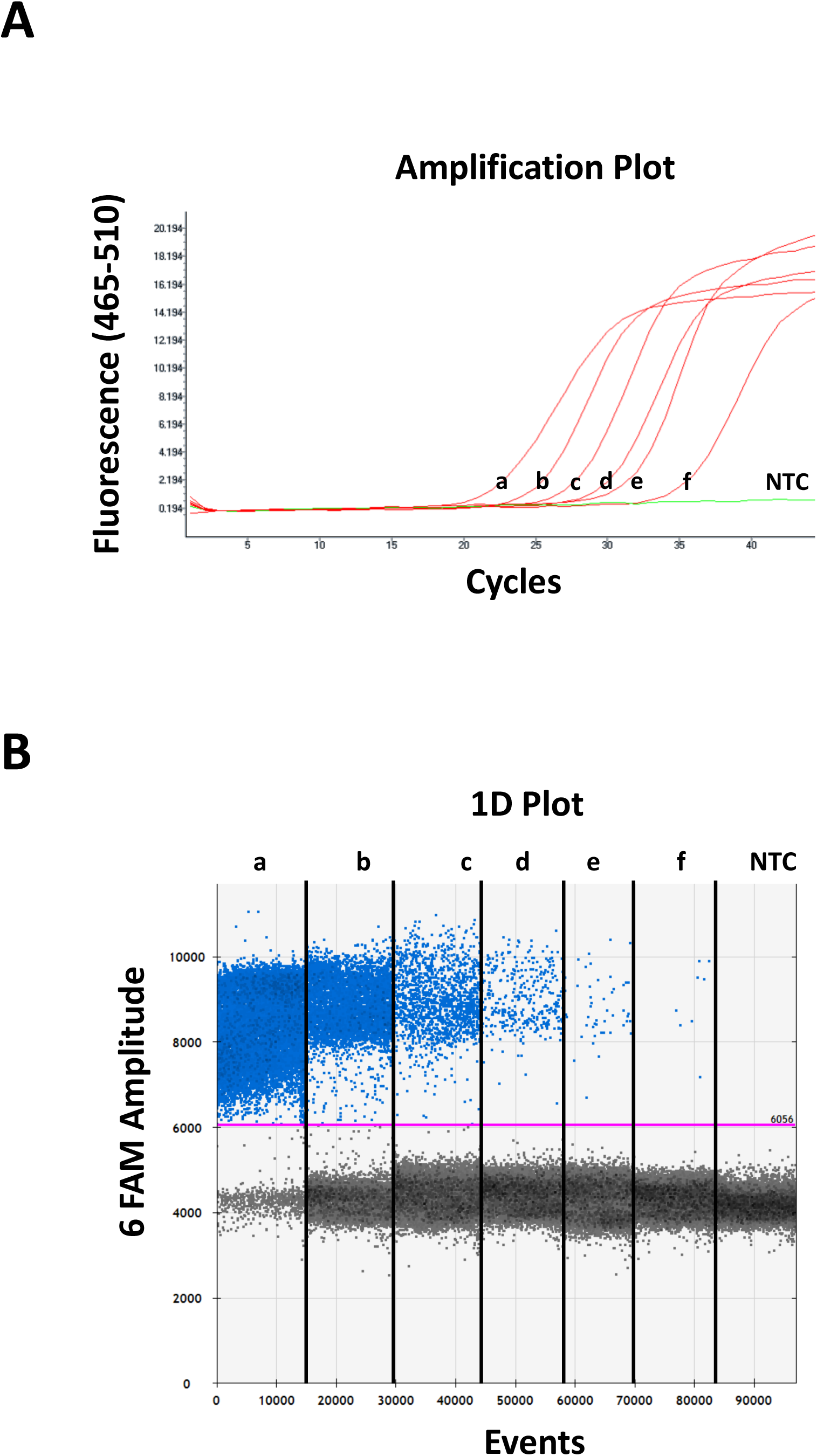
Serial dilution of viral RNA isolated with the in-house method. 5-fold serial dilutions of a patient’s sample isolated with the novel custom protocol (a-f). NTC was used to test sensitivity of both RT-qPCR (A) and RT-ddPCR (B).

These data allowed us to optimize the pretreatment protocol published by Fomsgaard and Rosenstierne^6^.

In conclusion, our treatment enables a SARS CoV-2 RNA amplification which is at least as sensitive and accurate as that performed with RNAs extracted using commercial kits. At this time of rapid spreading of COVID-19, the shortage of reagents for RNA extraction is significantly slowing down the testing process, which is the most effective way to contrast the pandemic. Our simple and fast protocol overcomes these limitations, bypassing the need for RNA extraction reagents, shortening the time of the analysis and reducing its cost.

## Data Availability

Data that support this study are available from the corresponding author upon reasonable request.

